# Development of serological assays to identify *Helicobacter suis* and *Helicobacter pylori* infections

**DOI:** 10.1101/2022.10.10.22280809

**Authors:** Hidenori Matsui, Emiko Rimbara, Masato Suzuki, Kengo Tokunaga, Hidekazu Suzuki, Masaya Sano, Takashi Ueda, Hitoshi Tsugawa, Sohachi Nanjo, Akira Takeda, Makoto Sasaki, Shuichi Terao, Tsuyoshi Suda, Sae Aoki, Keigo Shibayama, Hiroyoshi Ota, Katsuhiro Mabe

## Abstract

*Helicobacter suis* hosted by hogs is the most prevalent gastric non-*Helicobacter pylori Helicobacter* species found in humans. Recent studies suggest that the *H. suis* infection has already induced many cases of gastric disease. However, the infection period and route of *H. suis* from hogs remain unclear. Because diagnostic methods based on the urease activity of *H. suis* often yield negative judgments, there is no reliable method for diagnosing *H. suis* infection in clinical practice without gastric biopsy specimens. We developed the world’s first ELISA to simultaneously diagnose *H. suis* and *H. pylori* infection in a single test. The area under the ROC curve was 0.9648 or 0.9200 for identifying *H. suis* or *H. pylori* infection, respectively. The sensitivity, specificity, and positive and negative predictive values for identifying *H. suis* infection were 100%, 92.6%, 76.9%, and 100%, and those for identifying *H. pylori* infection were 88.2%, 87.5%, 65.2%, and 96.6%, respectively. (150 words)

## INTRODUCTION

*Helicobacter suis*, which differs from *H. pylori* in its bipolar flagella and corkscrew-like spiral morphology, is classified as a member of the gastric non-*H. pylori Helicobacter* species (NHPH), a group of bacteria that have been known to infect the human stomach since the 1980s^1-4^. The natural hosts of *H. suis* are macaques and hogs, while other NHPH such as *H. heilmannii* sensu stricto (s.s.), *H. ailurogastricus, H. felis, H. salomonis, H. baculiformis, H. cynogastricus*, and *H. bizzozeronii* infect the stomachs of cats and dogs^5-7^. Comparative genomic assessment of multilocus sequencing typing (MLST) of *H. suis* based on seven housekeeping genes revealed high genomic homology between hog- and human-derived isolates, suggesting that *H. suis* infection in humans is a zoonosis originating from hogs^5,8,9^.

Because the conventional *H. pylori* diagnostic tests based on the urease activity, such as the urea breath test (UBT) and rapid urease test (RUT), often yield negative judgments in the case of NHPH infecting the human stomach, PCR or histological examination from gastric biopsy specimens has been performed to diagnose NHPH infection for research purposes. In past reports, the prevalence of NHPH infection was shown to be less than 0.5% by microscopic detection of the NHPH morphology in gastric sections^10-12^. In more recent reports, Tsukadaira *et al*. estimated the NHPH infection rate in Japan to be approximately 3.35%^13^. When a PCR assay targeting urease genes was performed on the 30 patients with NHPH-associated gastritis, 28 were infected with NHPH as follows (urease genes were not detected in 2 patients): 26 with *H. suis* and 2 with *H. heilmannii* s.s. or *H. ailurogastricus*^13^. They concluded that *H. suis* was the most common NHPH that infected human stomachs^13^. The *H. suis* infection leads to diseases such as gastric mucosa-associated lymphoid tissue (MALT) lymphoma, nodular gastritis, chronic gastritis, and peptic ulcer^14-20^. However, the above reports did not provide proof of *H. suis* or other NHPH infection because they only used PCR assay and did not isolate the bacteria or analyze the isolated bacteria’s genome to identify the infecting bacteria.

Because most *H. pylori* infections occur in infancy, usually via transmission from family members, the infection rate of *H. pylori* has dramatically declined mainly due to improvements in the general hygiene environment in childhood in Japan^21^. As a result, the prevalence of *H. suis* infection could become more significant than the *H. pylori* infection rate if no measures are taken to prevent *H. suis* infection^22^. Because eradication therapy of *H. suis* is effective in *H. pylori*-negative gastric MALT lymphoma^17,23^, the *H. suis* infection could be a predictive marker for the efficacy of eradication therapy for *H. pylori*-negative gastric MALT lymphoma^24^. Therefore, the development of comprehensive methods to diagnose *H. suis* infection has become a matter of urgency for clinical practice.

Nonetheless, given the current worldwide infection status—i.e., more than half of the world’s population is still infected with *H. pylori*^25^—there is a need for rapid diagnostic products that can identify both *H. suis* and *H. pylori* infection simultaneously. This is the first study to report a method for the simultaneous serological diagnosis of *H. suis* and *H. pylori* infection in human subjects; the study also reports the evaluation of the anti-*H. suis* antibody titers after eradication.

## RESULTS

### Classification of the specimens

As shown in Table 1, by using routine methods, PCR, and culture for diagnosing *Helicobacter* infections, we classified all 101 subjects into the following four groups in advance: an *H. suis*-infection group (n=20), an *H. pylori*-infection group (n=17), a group with eradication of *H. pylori* infection (n=20), and a group with neither *H. suis* nor *H. pylori* infection (n=44). The patients in these groups had gastritis (n=52), gastric mucosa-associated lymphoid tissue (MALT) lymphoma (n=23), duodenal MALT lymphoma (n=1), peptic ulcer (n=20), autoimmune (metaplastic atrophic) gastritis (n=4), and endoscopic mucosal resection (EMR) for early gastric cancer (EGC; n=1). The percentage of men in the *H. suis* infection group was 85%, which was significantly different from the percentages in the *H. suis* non-infection groups, i.e., the *H. pylori* infection group (58.8%), the post-eradication of *H. pylori* infection group (45.0%), and the non-infection group (45.0%) (all *P*<0.05). Table S1 shows the individual data of the 101 examinees. The partial 23S rRNA gene sequences of the 13 *H. suis* and 16 *H. pylori* strains isolated in this study were compared with those of the reference *Helicobacter* strains. The genomic analysis successfully classified every *Helicobacter* strain with species specificity. All *H. suis* and *H. pylori* strains belonged to the same phylogenetic clades of *H. suis* and *H. pylori* reference strains, indicating that the genomic identification in this study was correct. We proved that all *H. suis* strains have more than 99% average nucleotide identity (ANI) values against the *H. suis* type strain HS1 (Figure S1).

**Table 1.**
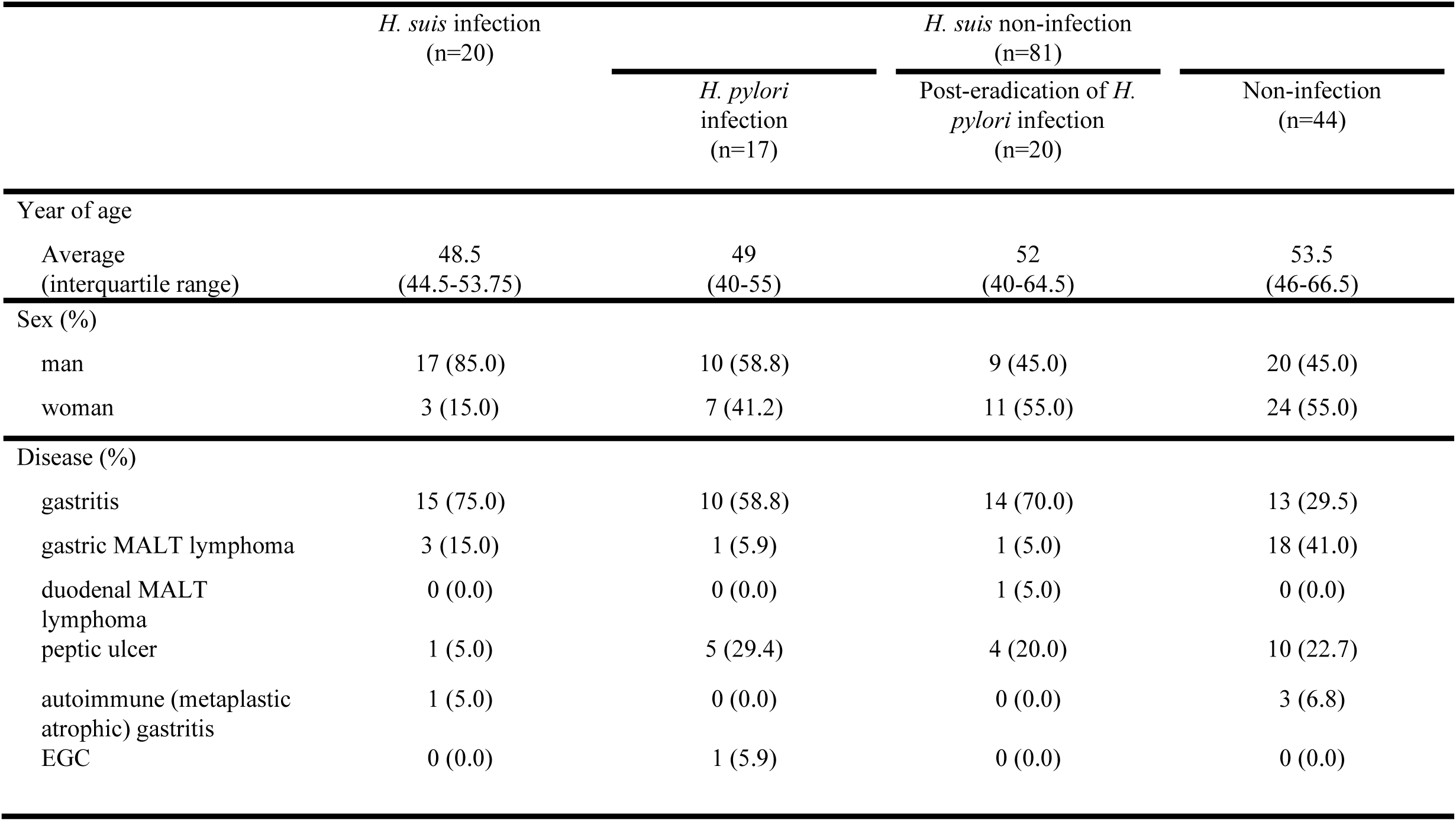
Examinee background characteristics.

### Measurement of the anti-*H. suis* and anti*-H. pylori* antibody titers

As shown in Figure 1, the mean with standard deviation (SD) of the absorbance at 450 nm of ELISA for identifying *H. suis* infection in the *H. suis* infection group was 2.124±0.535, which was significantly higher than those of the *H. suis* non-infection groups—i.e., the *H. pylori* infection group (0.530±0.807, *P*<0.0001), the post-eradication of *H. pylori* infection group (0.392±0.514, *P*<0.0001), and the non-infection group (0.261±0.447, *P*<0.0001) (Figure 1A). The cut-off value (0.997) for the absorbance at 450 nm of *H. suis* infection was calculated by receiver operating characteristic (ROC) curve analysis between the *H. suis* infection group (n=20) and the *H. suis* non-infection groups (n=81) —i.e., the *H. pylori* infection (n=17), post-eradication of *H. pylori* infection (n=20), and non-infection with either *H. suis* or *H. pylori* (n=44) groups (Figure 1B). Meanwhile, the mean with SD of the absorbance at 450 nm of ELISA for identifying *H. pylori* infection in the *H. pylor*i infection group was 1.372±0.715, which was significantly higher than those of two of the *H. pylori* non-infection groups—i.e., the *H. suis* infection group (0.458±0.441, *P*<0.0001) and the non-infection group (0.300±0.429, *P*<0.0001)—but not the post-eradication of *H. pylori* infection group (0.850±0.831, *P*>0.05) (Figure 1C). Then, the cut-off value (0.811) for the absorbance at 450 nm of *H. pylori* infection was calculated by ROC curve analysis between the *H. pylori* infection group (n=17) and the *H. pylori* non-infection groups (n=64), which were the *H. suis* infection group (n=20) and the group with non-infection of either *H. suis* or *H. pylori* (n=44) (Figure 1D). The sensitivity, specificity, and positive and negative predictive values (PPV and NPV, respectively) of ELISA for identifying the *H. suis* infection group against the *H. suis* non-infection groups were 100% (95% confidence interval [CI]: 83.9% to 100%), 92.6% (95% CI: 84.8% to 96.6%), 76.9% (95% CI: 55.9% to 90.2%), and 100% (95% CI: 93.9% to 100%), respectively (Table 2). On the other hand, the sensitivity, specificity, PPV, and NPV of ELISA for identifying the *H. pylori* infection group against the *H. pylori* non-infection groups were 88.2% (95% CI: 65.7% to 97.9%), 87.5% (95% CI: 77.2% to 93.5%), 65.2% (95% CI: 42.8% to 82.8%) and 96.6% (95% CI: 87.0% to 99.4%), respectively (Table 3).

**Table 2.** ELISA to identify *H. suis* infection. Positive includes the *H. suis* infection group (n=20). Negative includes the *H. suis* non-infection groups (n=81). All specimens (n=101) were classified into two groups: a group with an *H. suis*-infection value greater than or equal to the cut-off value (n=26) and a group with an *H. suis*-infection value smaller than the cut-off value (n=75). Cut-off value, 0.997; sensitivity, 20/(20+0)=100%; specificity, 75/(6+75)≈92.6%; PPV, 20/(20+6)≈76.9; NPV, 75/(0+75)=100%.

|  |  | Positive | Negative | Total |
| --- | --- | --- | --- | --- |
| Absorbance at 450 nm | $\geq 0.997$ | 20 | 6 | 26 |
| | $< 0.997$ | 0 | 75 | 75 |
|  | Total | 20 | 81 | 101 |

**Table 3.** ELISA to identify *H. pylori* infection. Positive includes the *H. pylori* infection group (n=17). Negative includes the *H. suis* infection group and the non-infection group (n=64). All specimens (n=81) were classified into two groups: a group with an *H. pylori*-infection value greater than or equal to the cut-off value (n=23) and a group with an *H. pylori*-infection value smaller than the cut-off value (n=58). Cut-off value, 0.811; sensitivity, 15/(15+2)≈88.2%; specificity, 56/(8+56)≈87.5%; PPV, 15/(15+8)≈65.2%; NPV, 56/(2+56)≈96.6%.

|  |  | Positive | Negative | Total |
| --- | --- | --- | --- | --- |
| Absorbance at 450 nm | $\geq 0.811$ | 15 | 8 | 23 |
| | $< 0.811$ | 2 | 56 | 58 |
|  | Total | 17 | 64 | 81 |

**Figure 1.**
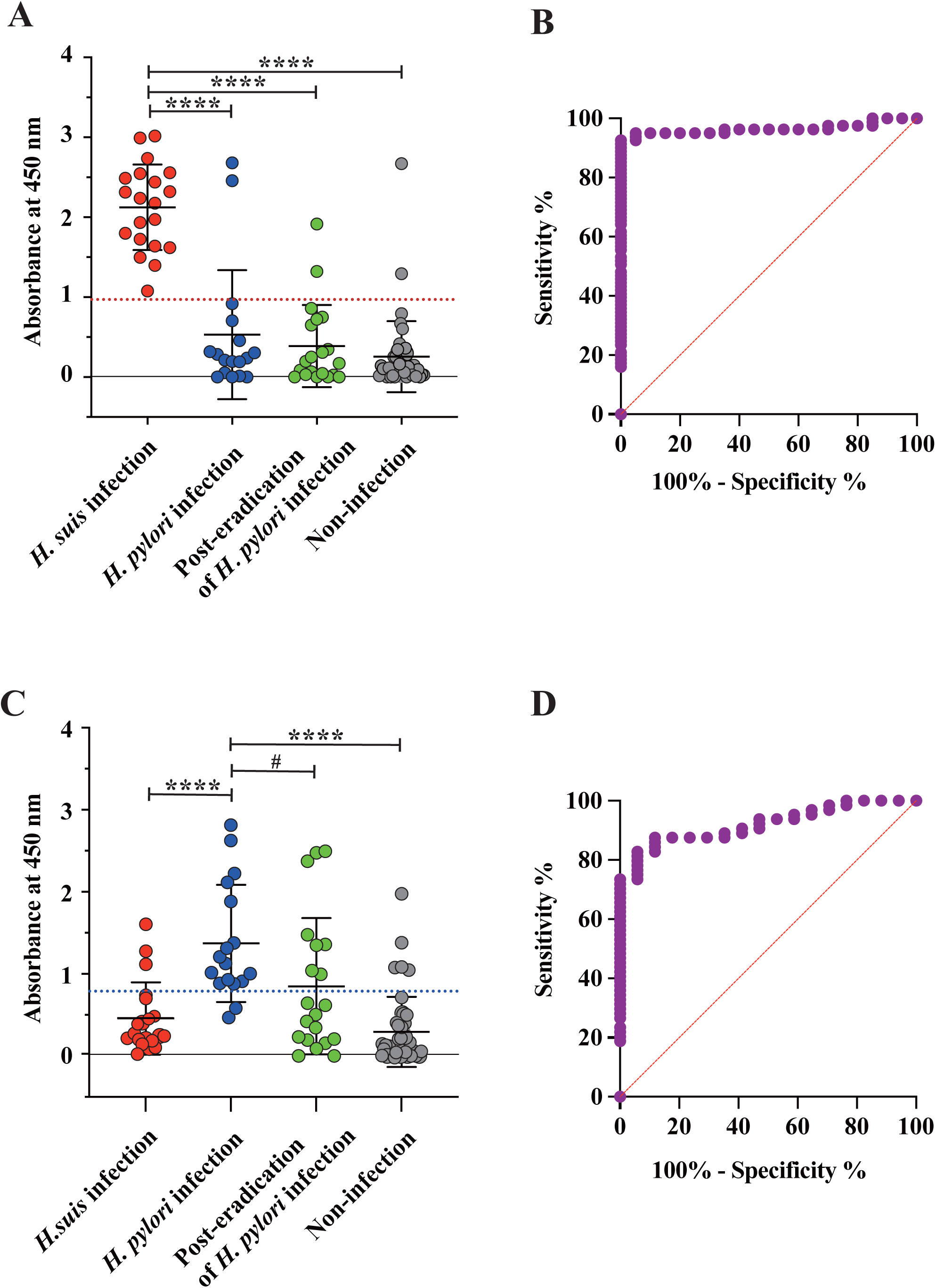
ELISA for identification of *H. suis* and *H. pylori* infection. Serum specimens (n=101) were divided into four groups: an *H. suis*-infection group (n=20), an *H. pylori*-infection group (n=17), a group with eradication of *H. pylori* infection (n=20), and a group with neither *H. suis* nor *H. pylori* infection (n=44). (a) ELISA for detecting *H. suis* infection. \*\*\**P*<0.0001, the *H. suis* infection group vs. the *H. pylori* infection, the post-eradication of *H. pylori* infection, or the non-infection group. The red dotted line indicates the cut-off value (0.997). The bar represents the mean with SD. (b) ROC curve constructed from the *H. suis*-infection group vs. the *H. suis* non-infection groups, which consisted of the *H. pylori* infection, the post-eradication of *H. pylori* infection, and the non-infection group. The red slanted line is the reference line. The area under the ROC curve (AUC): 0.9648±0.01794; 95% CI: 0.9297-1.000; *P*<0.0001. (c) ELISA for detecting *H. pylori* infection. \*\*\*\**P*<0.0001, the *H. pylori* infection group vs. the *H. suis* infection or non-infection group. #*P*>0.05, the *H. pylori* infection vs. the post-eradication of *H. pylori* infection group. The blue dotted line indicates the cut-off value (0.811). The bar represents the mean with SD. (d) ROC curve constructed from the *H. pylori* infection group vs. the *H. pylori* non-infection groups, including the *H. suis* infection and the non-infection groups. The red slanted line is the reference line. AUC: 0.9200±0.02957; 95% CI: 0.8621-0.9780; *P*<0.0001.

### Decline of anti-*H. suis* antibody titers after eradication

Among 20 individuals belonging to the *H. suis* infection group, 6 individuals (nos. 94058, 94059, 04080, 04252, 04262, and 14005) suffering from gastritis were administered the eradication therapy as described in the Methods section. After the eradication therapies of *H. suis*, the upper gastrointestinal (GI) endoscopic pathology and PCR for the detection of *H. suis* were performed, and successful eradication of *H. suis* infection was judged in all 6 of these patients. The anti-*H. suis* antibody titers decreased over time after eradication. The estimated times required for the anti-*H. suis* antibody titers to decline to 0.997 (the cut-off value for the absorbance at 450 nm) in the 6 specimens were as follows: 15.1 months (no. 94058), 12.3 months (no. 94059), 9.1 months (no. 04080), 1.6 months (no. 04252), 3.1 months (no. 04262), and 2.8 months (no. 14005). Then, the 6 individuals were divided into the following two groups: a group of individuals exhibiting a slow decline of anti-*H. suis* antibody titers after eradication (nos. 94058,94059, and 04080) and a group exhibiting a fast decline of anti-*H. suis* antibody titers after eradication (nos. 04252, 04262, and 14005). The time required for the anti-*H. suis* antibody titers to decline until the cut-off value was re-calculated at 13.6 and 2.6 months after eradication in the slow (Figure 2A) and first (Figure 2B) decline groups, respectively.

**Figure 2.**
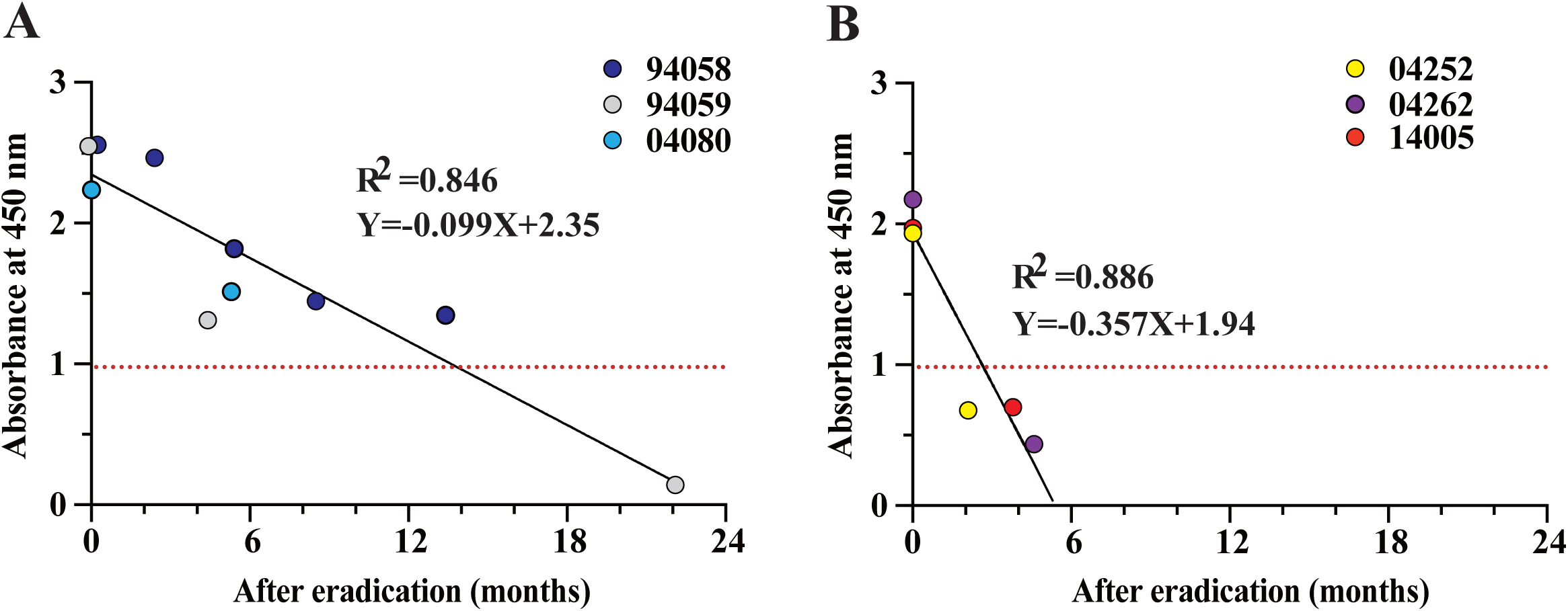
Measurement of anti-*H. suis* antibody titers after the eradication of *H. suis* infection. Six individuals underwent *H. suis*-eradication treatment, and then were monitored for serum anti-*H. suis* antibody titers by ELISA. Results for 3 individuals (nos. 94058, 94059,and 04080) (A) were plotted separately from the other 3 (nos. 04252, 04262, and 14005) (B). The red dotted line indicates the cut-off value.

## DISCUSSION

### The infection source of *H. suis*

Twenty out of 101 examinees were infected with *H. suis*, but no other NHPH were identified by the culture and PCR (negative and positive judgments in PCR targeting the *hsvA* and NHPH 16S rRNA genes, respectively). Therefore, we presume a lower risk of infection with NHPH in the human stomach other than *H. suis* to be colonizing in the stomachs of cats and dogs. On the other hand, no *H. suis* infection has yet been identified in the stomachs of cats and dogs^5-7,26,27^. Although *H. suis* colonizes the hog stomach and is the source of zoonotic infection in a high percentage of human cases of *H. suis* infection^28-30^, the infection rate among hog farmers is not higher than that of individuals with no contact with animals^31,32^. It has been hypothesized that *H. suis* infects humans via the consumption of contaminated pork^33,34^. However, there is no clear evidence regarding the *H. suis* infection route or period. In the present study, we did not find a mixed infection with *H. pylori* and *H. suis*.

### Validation of the serological assay for the detection of *H. suis* and *H. pylori* infection

ELISA was performed by a separate institute using the same protocol and serum from a separate container. The data obtained were consistent with those obtained in every laboratory. The obtained cut-off values in this study can be reproduced for studies in other institutes. The ELISA has always been verified using 1800-fold diluted serum in addition to the 3600-fold diluted serum (Figure S2). Positive and negative judgments in both trials have been consistent. Marini *et al*. have tried detecting anti-*Helicobacter* antibodies of macaques by ELISA using the *H. pylori* SS1 and *H. suis* outer membrane protein preparations as antigens^35^. In the identification of *H. suis* infection, the sensitivity and specificity were 70.0% (95% CI: 34.8% to 93.3%) and 75.0% (95% CI: 19.4% to 99.4%), respectively^35^. Meanwhile, in the identification of *H. pylori* infection, the sensitivity and specificity were 66.7% (95% CI: 9.2% to 99.2%) and 63.6% (95% CI: 30.8% to 89.1%), respectively^35^. Therefore, the ELISA data using the whole bacterial cell lysates as antigens and the 3600-fold diluted sera in this study (Figure 1 and Tables 2 and 3) showed more accuracy than the ELISA data using the outer membrane protein preparations as antigens. When synthetic peptides constructed from each *H. suis* or *H. pylori*-specific autotransporter protein sequence were used as antigens, ELISA did not show higher accuracy compared to whole-cell lysates as antigens.

### Discrimination of the judgments between the serological assay and PCR or culture

We conducted conventional Giemsa staining and immunohistochemistry (IHC) to identify *H. suis* and *H. pylori* in gastric biopsy specimens. As shown in Figure S3, *H. suis* was detected in the lumen of foveola and on the epithelial cells by the IHC test using the rabbit monoclonal anti-*H. pylori* antibody (clone EP279) (Figure S3B) or the rabbit polyclonal anti-*H. suis* antibody (Figure S3C) in the no. 04029 biopsy specimen (*H. suis* infection group). In contrast, the IHC test using the rabbit monoclonal anti-*H. pylori* antibody detected *H. pylori* (Figure S3E), and the rabbit polyclonal anti-*H. suis* antibody weakly detected *H. pylori* (Figure S3F) in the no. 04220 biopsy specimen (*H. pylori* infection group). These results indicate that although the rabbit polyclonal anti-*H. suis* antibody reacts explicitly with *H. suis*, the rabbit monoclonal anti-*H. pylori* antibody exhibited cross-reactivity with *H. suis*. In contrast, Giemsa staining of biopsy specimens could clearly distinguish spiral forms of *H. suis* (Figure S3A) from spiral forms of *H. pylori* (Figure S3D). To compare the discriminatory ability between the serological assay and PCR or culture using gastric biopsy specimens, we attempted to detect *H. suis* and *H. pylori* by Giemsa staining and IHC analysis of the biopsy specimens. Among 17 (*H. pylori* infection group) and 20 (post-eradication of *H. pylori* infection group) individuals, 4 serum specimens (nos. 04232 and 04133 of *H. pylori* infection group, and nos. 04150 and 04162 of post-eradication of *H. pylori* infection group) were *H. suis*-positive by ELISA (Table S1). Because the microscopy did not detect *H. suis* in these specimens, we suggest that the whole-cell lysate of *H. suis* SNTW101c reacted with anti-*H. pylori* serum antibodies. Among 44 individuals in the non-infection group, 2 serum specimens (nos. 04293 and 04018 from patients with gastric MALT lymphoma and peptic ulcer, respectively) were *H. suis*-positive by ELISA (Table S1). However, the microscopy did not detect *H. pylori* or *H. suis*. We presume that the individuals had been infected with *H. suis* (no. 04293) and *H. pylori* (no. 04018) in the past because the serum specimens were clearly positive by ELISA for *H. suis* infection (no. 04293) and *H. pylori* infection (no. 04018) (Table S1). Moreover, in the non-infection group, we presume that the other 4 individuals had been infected in the past with *H. pylori* and the *H. pylori* was spontaneously eradicated because the serum specimens (nos. 04167, 14327, 04157, and 14202) were *H. pylori*-positive by ELISA (Table S1).

### Evaluation of the *H. suis* eradication therapies by serological assay

When *H. suis* infection becomes apparent, eradication of *H. suis* is recommended even if the examinees are not suffering from severe illnesses such as gastric MALT lymphoma, nodular gastritis, or intractable peptic ulcer. The regimen for *H. pylori* eradication has been used for NHPH eradication. In most cases, patients are treated with triple-agent eradication therapy consisting of an inhibitor of gastric acid secretion such as a PPI or a potassium-competitive acid blocker (P-CAB) along with two antibiotics such as amoxicillin and clarithromycin or amoxicillin and metronidazole for 7 to 14 days^14,15,18,36-38^. In the current study, the time-dependent decline of antibody titers after eradication confirmed the success of the eradication therapies. There were two patterns of the decline of anti-*H. suis* antibody titers—namely, a faster or a slower decline after eradication therapies (Figure 2). The difference in eradication efficacy between the slow and fast decline groups seems to be attributed to the difference in antibody titers before eradication (2.446±0.181 and 2.027±0.130 in the slow and fast decline groups, respectively, *P*<0.05). The serological assay of *H. suis* infection would be beneficial as a follow-up examination to confirm eradication (Figure 2).

### The urgent requirements

*H. pylori* infection in infancy might confer years of protection against *H. suis* infection. In the present study, none of the *H. suis* infected examinees had *H. pylori* infection or prior *H. pylori* infection that had been eradicated. The *H. suis* infection rate might have risen with the decline of the *H. pylori* infection rate. Because there is currently no established method for the clinical diagnosis of *H. suis* infection, accurate diagnosis of *H. suis* infection is crucial in order to avoid wasting precious time on the wrong course of treatment. A serological assay to simultaneously identify both *H. suis* and *H. pylori* infection is available for the large-scale periodic medical examination of unimpaired individuals; this assay can provide an accurate estimation of the infection rate and can effectively prevent gastropathy.

## Supporting information

Supplemental method, Figure S1, S2, andS3

## Data Availability

All data produced in the present work are contained in the manuscript.

## LIMITATION OF THE STUDY

This study is the first to develop a serological diagnostic method for *H. suis* infection in humans. The sensitivity, specificity, PPV, and NPV of ELISA for identifying the *H. suis* infection group against the *H. suis* non-infection groups were 100% (95% CI: 83.9% to 100%), 92.6% (95% CI: 84.8% to 96.6%), 76.9% (95% CI: 55.9% to 90.2%), and 100% (95% CI: 93.9% to 100%), respectively. On the other hand, the sensitivity, specificity, PPV, and NPV of ELISA for identifying the *H. pylori* infection group against the *H. pylori* non-infection groups were 88.2% (95% CI: 65.7% to 97.9%), 87.5% (95% CI: 77.2% to 93.5%), 65.2% (95% CI: 42.8% to 82.8%) and 96.6% (95% CI: 87.0% to 99.4%), respectively. With respect to diagnostic accuracy, the low values of PPV were the limitation of this serological assay. In cases with a negative judgement by PCR, previously eradicated infections might lead to a positive judgement by ELISA.

## SUPPLEMENTAL INFORMATION

Supplemental information is attached separately from the main text as Excel (Table S1) and PDF (Figures S1, S2, and S3) files.

## ACKNOWLEDGMENTS

This work was supported by MEXT/JSPS KAKENHI under grant numbers JP19H03474 (H.M.) and JP20K08365 (T.K.), and by AMED under grant numbers JP20fk0108148 (E.R.) and JP20fk0108133 (M.S.).

## AUTHOR CONTRIBUTIONS

H.M., E.R., and K.S. conceptualized the research. E.R. and S.A. performed the cultures and PCR. M.S. performed the genomic analysis. H.M. performed ELISA. H.O. performed microscopy. K.M. and K.T. performed the *H. suis*-eradication treatment. K.M., K.T., H.S., M.S., T.U., H.T., S.N., A.T., M.S., S.T., and T.S. collected specimens. H.M., E.R., and K.M. prepared the manuscript. All authors verified the underlying data and approved the final version of the manuscript for publication.

## STAR METHODS

### KEY RESOURCES TABLE

Attached separately from the main text.

#### Lead contact

Further information and requests for resources and reagents should be directed to and will be fulfilled by the lead contact, Hidenori Matsui

#### Materials availability

The polyclonal anti-*H. suis* antibody and ELISA prototype generated by this study are available from the lead contact upon reasonable request. Bacterial strains isolated in this study are also available from the lead contact upon reasonable request.

#### Data and code availability

The *H. suis* whole-genome data and the *H. pylori* 23S rRNA genome data have been deposited at GenBank/ENA/DDBJ and will be available from the date of publication. Accession numbers are listed in the Tables S1. Any additional information about the analysis in this paper is available from the lead contact upon reasonable request.

## METHODS DETAILS

### Ethics statement

The Research Ethics Committees (REC) of the National Institute of Infectious Diseases (NIID) and Kitasato University approved this study under registration numbers 1284 and 18100, respectively. The ethical review was also approved at the following eight medical institutions: Junpukai Health Maintenance Center, Kyorin University Hospital, Tokai University Hospital, Toyama University Hospital, Takeda Hospital, Aichi Medical University Hospital, Kakogawa Central Municipal Hospital, and Kanazawa Municipal Hospital. We also obtained written informed consent from all examinees, each of whom received an explanation of the study at one of the participating medical institutions.

### Collection and transportation of clinical specimens

Serum and gastric biopsy specimens from examinees undergoing medical check-ups or suffering from any gastric disorder who underwent upper GI endoscopy at one of the eight above-named medical institutions were enrolled. However, the examinees with normal stomachs and those with a gastric disease under 20 years of age were excluded from this study. The specimens from participants having healthy stomachs were excluded from this study. All examinees were found to have *H. pylori* infection by means of two of the following five tests: the UBT, RUT, serum *H. pylori* antibody test, stool *H. pylori* antigen test, and histological test. *H. pylori, H. suis*, and other NHPH infections were finally confirmed by culture of gastric biopsy specimens. Gastric biopsy specimens were collected from the greater curvature of the gastric antrum, the greater curvature of the lower gastric body, and the lesser curvature of the gastric angle, and they were transported to the NIID using the NHPH biopsy transport media for cultivation, PCR, and microscopy. Serum specimens were divided into three containers, transported to the NIID, and stored at -80°C until use. The specimen number assigned by the NIID and the clinical data cannot be used to identify medical institutions or examinees.

### Culture

The gastric biopsy specimen was homogenized with 300 μL of Brucella broth (BD, Franklin Lakes, NJ, USA) adjusted to pH 5.0 by hydrochloride. A 200 μL aliquot of the tissue homogenates was inoculated onto NHPH agar plates containing 1.5% (wt/vol) agar, Brucella broth, 20% (vol/vol) heat-inactivated bovine serum (FBS), *Campylobacter* selective supplement (Skirrow; Oxoid, Basingstoke, UK), Vitox supplement (Oxoid), and hydrochloric acid to adjust the pH to 5.0, and incubated for more than seven days in a humidified gas mixture (5% O_2_, 12% CO_2_, and 83% N_2_) at 37°C. The grown colonies of primary culture were inoculated onto NHPH agar plates and enriched by modified biphasic culture for 120 h with shaking in a humidified gas mixture at 37°C. The whole-genome sequencing of broth-grown bacteria was carried out as described below. Although both *H. pylori* and NHPH were grown on the NHPH agar plate as primary cultures, the *H. pylori* colonies tended to grow faster and to be larger than the small, slow-growing colonies formed by NHPH^9^. Therefore, the colonies formed by *H. pylori* were inoculated onto a Brucella agar plate containing 5% (vol/vol) horse blood for 72 h in a humidified gas mixture at 37°C. NHPH could not grow on this plate.

### PCR

To identify NHPH infections, the DNA was prepared from the remaining 100 μL of homogenates of gastric biopsy specimens using DNeasy Blood & Tissue Kits (Qiagen, Hilden, Germany). Then, the DNA was used as the template for probe-based real-time PCR targeting of the *H. suis*-specific *vacA*-type autotransporter protein gene (*hsvA*) and the NHPH-common region of the 16S rRNA gene. The PCR procedure was described previously^9^. The sequences of the two sets of primers and probes were as follows: NHP194003_11930_forward (5’-CTGGTAATGCATCATTAGAAGCAAA-3’), NHP194003_11930_reverse (5’-GATGGGCGCTTCTGGTTTA-3’), and NHP194003_11930_probe (5’-/56-FAM/TGTACACAC/ZEN/CAAACAGATGAGCCGT/3IABkFQ-3’) for targeting the *hsvA* gene; NHPH_16S_F (5’-CAAGTCGAACGATGAAGCCTA-3’), NHPH_16S_R (5’-ATTTGGTATTAATCACCATTTCTAGT-3’), and NHPH_16S_probe (5’-/56-FAM/TTACTCACC/ZEN/CGTGCGCCACTAATC/3IABkFQ/-3’) for targeting the NHPH 16S rRNA gene. The diagnosis of *H. suis* infection was made when both positive results occurred in two types of PCR targeting the *hsvA* and NHPH 16S rRNA genes^9^. The diagnosis of NHPH other than *H. suis* was made when negative and positive results occurred in PCR targeting the *H. suis*-specific *hsvA* and NHPH-common 16S rRNA genes, respectively. We included the culture as a diagnostic criterion to confirm *H. pylori* infection, whereas we excluded the culture as a diagnostic criterion to confirm *H. suis* infection. To identify *H. pylori* infection, the 651 bp DNA fragment of the 23S rRNA gene was amplified by the colony PCR using the following set of primers: F3 (5’-CCGTAGCGAAAGCGAGTCT-3’) and R3 (5’-CCCGACTAACCCTACGATGA-3’). The 23S rRNA gene sequences of *Helicobacter pylori* strains were deposited at GenBank/EMBL/DDBJ as shown in Table S1. The nucleotide sequences of PCR products were aligned by ClustalW ver. 2.1 (http://www.clustal.org/clustal2/). The 23S rRNA gene sequences of the reference *Helicobacter* strains were taken from GenBank/EMBL/DDBJ.

### Broth culture of *H. pylori* and *H. suis* for the preparation of ELISA antigens

The colonies of *H. pylori* TN2GF4 grown on a *Helicobacter*-selective agar plate (Nissui Pharmaceutical, Tokyo) were inoculated into a Brucella broth containing 10% (vol/vol) FBS supplemented with 1% (vol/vol) Vitox supplement and *Helicobacter pylori-*selective supplement (Dent; Oxoid) for 72 h by shaking in a humidified gas mixture (5% O_2_, 10% CO_2_, and 85% N_2_) at 37°C^39^. The frozen stock of *H. suis* SNTW101c was inoculated onto NHPH agar plates and enriched by modified biphasic culture for 120 h with shaking in a humidified gas mixture at 37°C as described above.

## ELISA

Bacterial cells collected by centrifugation were washed twice with phosphate-buffered saline, pH 7.4 (PBS), suspended in distilled water and disrupted by sonication with a Bioruptor II (BMBio, Tokyo) to prepare whole-cell lysates. For the measurement of serum anti-*H. suis* or anti*-H. pylori* antibody titer, Nunc-Immuno 96-well microtiter plates (No. 439454; Thermo Fisher Scientific, Waltham, MA, USA) containing 100 μL of whole-cell lysates of *H. suis* SNTW101c or *H. pylori* TN2GF4 (4 μg/mL in 0.1 M carbonate/bicarbonate buffer, pH 9.4) were incubated overnight at 4°C. After washing three times with PBS containing 0.05% (vol/vol) Tween 20 (PBS-T), the wells were saturated with 200 μL of blocking buffer (PBS containing 1% BSA). The plates were incubated for 1 h at 37°C while shaking at 500 rpm. After washing three times with PBS-T, the wells were filled with 50 μL of serum samples diluted at 1:1,800 (Figure S21) and 1:3,600 (Figure 1) with blocking buffer. The plates were incubated for 1 h at 37°C while shaking at 500 rpm. After washing three times with PBS-T, the wells were filled with 50 μL of horseradish peroxidase (HRP)-conjugated goat anti-human IgA+IgG+IgM (H+L) secondary antibody (Jackson, Bar Harbor, ME, USA) diluted at 1:10,000 with blocking buffer. The plates were incubated for 1 h at 37°C while shaking at 500 rpm. After washing three times with PBS-T, the wells were filled with 50 μL of KPL SureBlue TMB Microwell Peroxidase Substrate (1-Component) (Sera Care Life Sciences, Milford, MA, USA). After 5 min, the color reaction was stopped by adding 50 μL of 1 N hydrochloric acid into the wells. The absorbance at 450 nm (reference wavelength, 630 nm) was measured on a ChroMate-6 microplate reader (Microtec, Chiba, Japan).

### Eradication of *H. suis*

For the treatment of gastric diseases with *H. suis* infection, each of the following three agents was administered twice daily for a week for eradication of *H. suis*: vonoprazan (P-CAB: 20 mg), amoxicillin (750 mg), and clarithromycin (200 mg for nos. 04080, 04252, 04262, and 14005 or 400 mg for nos. 94058 and 94059). At least two months after the eradication treatment, an upper GI endoscopy was performed to confirm gastric disease remission and the disinfection of *H. suis* by PCR.

### Statistics

Differences in the incidence between two groups and in the gender ratio among four groups were compared by Pearson’s chi-square and Fisher’s exact tests, respectively. The anti-*H. suis* and anti-*H. pylori* antibody values were compared among four groups using one-way ANOVA, followed by Tukey’s post hoc test. The linear regression lines were constructed by the least-squares method. Cut-off values, AUCs, sensitivities, and specificities based on the ROC curves were created using GraphPad Prism software version 9.5.0 for Mac (GraphPad Software, San Diego, CA, USA). As the diagnostic accuracy by ELIS, the PPV and NPV with 95% CI were calculated using Clinical Calculator 1 (http://vassarstats.net/clin1.html). Differences were considered significant when the *p-*value was less than 0.05.

