## Supplemental method, Figure S1, S2, andS3 for "Development of serological assays to identify *Helicobacter suis* and *Helicobacter pylori* infections"

[1]Department of Bacteriology II, National Institute of Infectious Diseases, Tokyo 208-0011, Japan
[2]Department of Bacteriology, Nagoya University Graduate School of Medicine, Aichi 466-8550, Japan
[3]Ōmura Satoshi Memorial Institute, Kitasato University, Tokyo 108-8641, Japan
[4]Antimicrobial Research Center, National Institute of Infectious Diseases, Tokyo 189-0002, Japan
[5]Department of General Medicine, Kyorin University School of Medicine, Tokyo 208-0011, Japan
[6]Division of Gastroenterology and Hepatology, Department of Internal Medicine, Tokai University School of Medicine, Kanagawa 259-1193, Japan
[7]Division of Host Defense Mechanism, Tokai University School of Medicine, Kanagawa 259-1193, Japan
[8]Third Department of Internal Medicine, Toyama University Hospital, Toyama 930-0194, Japan
[9]Takeda Hospital, Fukuoka 822-0027, Japan
[10]Division of Gastroenterology, Department of Internal Medicine, Aichi Medical University School of Medicine, Aichi 480-1103, Japan
[11]Department of Gastroenterology, Kakogawa Central City Hospital, Hyogo 675-8611, Japan
[12]Department of Gastroenterology, Kanazawa Municipal Hospital, Ishikawa 921-8105, Japan
[13]Department of Biomedical Laboratory Sciences, Shinshu University School of Medicine, Nagano 390-8621, Japan
[14]Junpukai Health Maintenance Center Kurashiki, Okayama 712-8014, Japan
[15]Lead contact

H. M. and E. R. contributed equally to this work.
*Corresponding authors at: Department of Bacteriology II, National Institute of Infectious Diseases, Gakuen 4-7-1, Musashimurayama-shi, Tokyo 208-0011, Japan and Junpukai Health Maintenance Center Kurashiki, Tsurashima 3-1-19, Kurashiki-shi, Okayama 712-8014, Japan. E-mail addresses: (to H. M.) and (to K. M.).

Figure S1

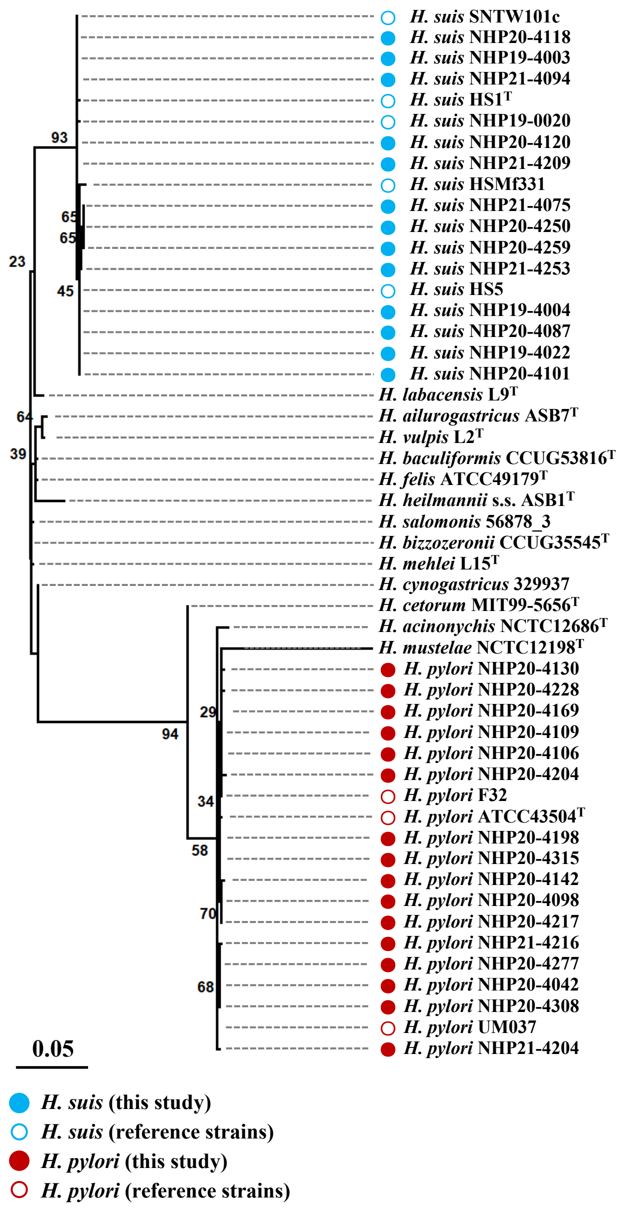
H. suis SNTW101c
H. suis NHP20-4118
H. suis NHP19-4003
H. suis NHP21-4094
H. suis HS1T
H. suis NHP19-0020
H. suis NHP20-4120
H. suis NHP21-4209
H. suis HSMf331
H. suis NHP21-4075
H. suis NHP20-4250
H. suis NHP20-4259
H. suis NHP21-4253
H. suis HS5
H. suis NHP19-4004
H. suis NHP20-4087
H. suis NHP19-4022
H. suis NHP20-4101
H. labacensis L9T
H. ailurogastricus ASB7T
H. vulpis L2T
H. baculiformis CCUG53816T
H. felis ATCC49179T
H. heilmannii s.s. ASB1T
H. salomonis 56878_3
H. bizzozeronii CCUG35545T
H. mehlei L15T
H. cynogastricus 329937
H. cetorum MIT99-5656T
H. acinonychis NCTC12686T
H. mustelae NCTC12198T
H. pylori NHP20-4130
H. pylori NHP20-4228
H. pylori NHP20-4169
H. pylori NHP20-4109
H. pylori NHP20-4106
H. pylori NHP20-4204
H. pylori F32
H. pylori ATCC43504T
H. pylori NHP20-4198
H. pylori NHP20-4315
H. pylori NHP20-4142
H. pylori NHP20-4098
H. pylori NHP20-4217
H. pylori NHP21-4216
H. pylori NHP20-4277
H. pylori NHP20-4042
H. pylori NHP20-4308
H. pylori UM037
H. pylori NHP21-4204
93
65
65
45
23
64
39
29
94
34
58
70
68
0.05
H. suis (this study)
H. suis (reference strains)
H. pylori (this study)
H. pylori (reference strains)

Figure S2

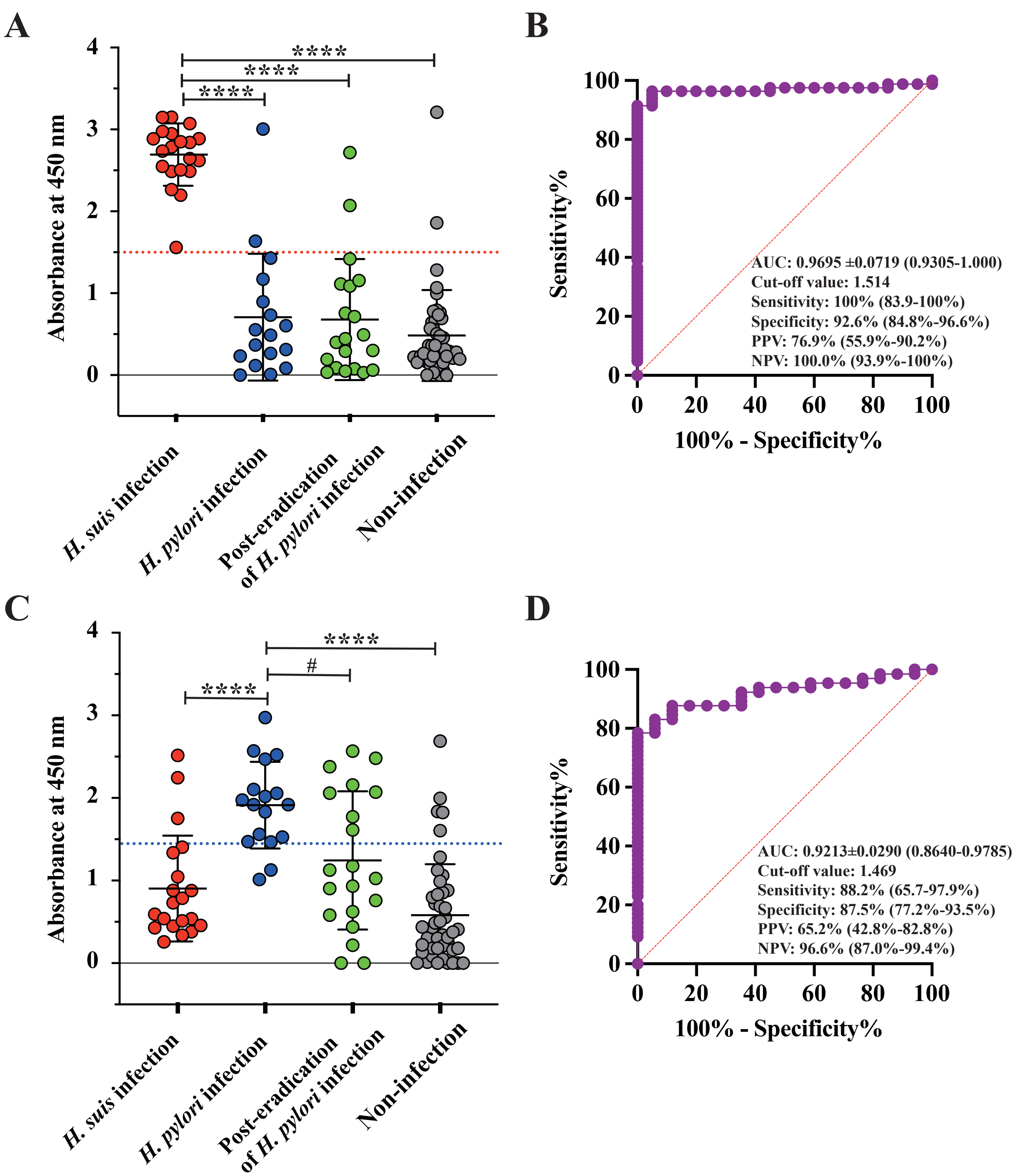

A
B
C
D
Absorbance at 450 nm
H. suis infection
H. pylori infection
Post-eradication of H. pylori infection
Non-infection
****
#
Sensitivity%
100% - Specificity%
AUC: 0.9695 ±0.0719 (0.9305-1.000)
Cut-off value: 1.514
Sensitivity: 100% (83.9-100%)
Specificity: 92.6% (84.8%-96.6%)
PPV: 76.9% (55.9%-90.2%)
NPV: 100.0% (93.9%-100%)
AUC: 0.9213±0.0290 (0.8640-0.9785)
Cut-off value: 1.469
Sensitivity: 88.2% (65.7-97.9%)
Specificity: 87.5% (77.2%-93.5%)
PPV: 65.2% (42.8%-82.8%)
NPV: 96.6% (87.0%-99.4%)

Figure S3

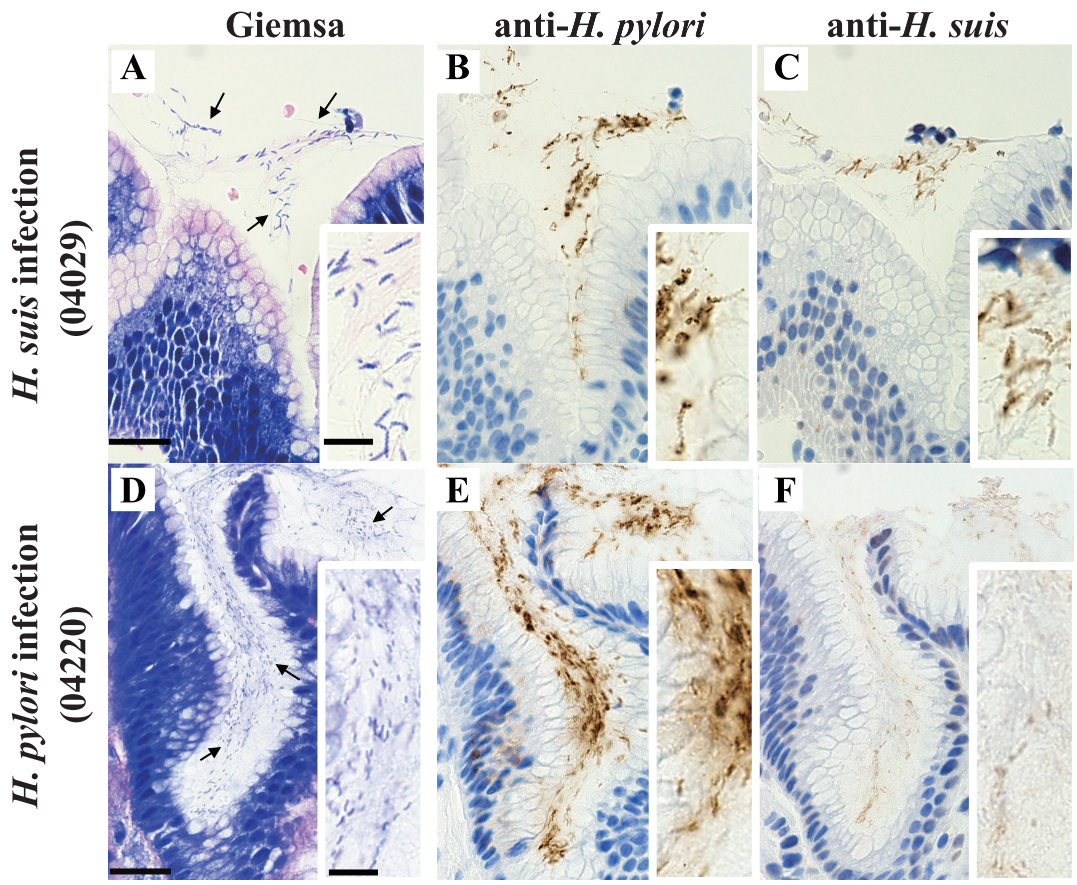

Giemsa
anti-*H. pylori*
anti-*H. suis*
*H. suis* infection (04029)
*H. pylori* infection (04220)
A
B
C
D
E
F

### SUPPLEMENTAL METHODS

#### Genomic analysis

Whole-genome sequencing of all culture-derived *H. suis* strains was performed using MiSeq (Illumina, San Diego, CA, USA). The library for Illumina sequencing (150-bp paired-end, insert size of 500-900 bp) was prepared using a Nextera XT DNA Library Prep Kit. The Illumina reads were assembled de novo using Shovill v1.1.0. (https://github.com/tseemann/shovill) with default parameters in order to acquire the draft genome sequences. Core genome alignments among *Helicobacter* strains were determined using Roary version 3.13.0 (https://github.com/sanger-pathogens/Roary) with default parameters. The maximum-likelihood phylogenetic trees were constructed by RAxML-NG v. 1.1 (https://github.com/amkozlov/raxml-ng) using partial 23S rRNA gene sequences or 628 core gene alignments. Whole-genome sequences of 15 strains of *H. suis* and 23S rRNA gene sequences of 16 strains of *H. pylori* were deposited at GenBank/EMBL/DDBJ as shown in Table S1. The bacterial species were determined by calculating the average ANI using pyani 0.2.12 (https://github.com/widdowquinn/pyani)[1]. MLST was performed according to the method described previously[1]. STs were assigned by using an online reference database (https://pubmlst.org/organisms/helicobacter-suis).

#### Microscopy

Polyclonal anti-*H. suis* antisera were prepared by immunization of two male New Zealand White rabbits with a bacterial lysate of heat-killed *H. suis* strain NHP20-4056, obtained from a 65-year-old man suffering from gastric MALT lymphoma. Polyclonal anti-*H. suis* IgG was purified using protein A Sepharose. Rabbit monoclonal anti-*H. pylori* antibody (clone EP279) was purchased from Cell Marque (Rocklin, CA, USA). For the histological study, 10% neutral-buffered, formalin-fixed, paraffin wax-embedded tissue blocks from gastric biopsy specimens were retrieved. Serial paraffin sections with a thickness of 3 μm were prepared. The sections were stained with Giemsa. For the IHC, the sections were deparaffinized, rehydrated, and placed in a 0.3% hydrogen peroxide solution in methanol for 30 min to block endogenous peroxidase activity. For the antigen retrieval, the sections in citrate buffer (pH 6.0) or Histofine antigen-retrieval solution (pH 9.0; Nichirei Biosciences, Tokyo) were heated at 110 °C for 10 min using a Decloaking Chamber NxGen (Biocare Medical, Pacheco, CA, USA). After blocking with 5% (wt/vol) BSA for 10 min, the sections were incubated with anti-*H. suis* IgG (13.2 µg/mL) or anti-*H. pylori* antibody (1:100 dilution) for 2 h, followed by rinsing three times in Tris-buffered saline (TBS, pH 7.4) and incubation with the HRP-conjugated goat anti-rabbit antibody (Agilent Technologies, Santa Clara, CA, USA; 1:50 dilution) for 1 h. The sections were stained with 0.03% (wt/vol) 3,3-diaminobenzidine tetrahydrochloride and 0.01% (vol/vol) hydrogen peroxide. Hematoxylin was used for counterstaining. The images were captured with a BX53 upright microscope (Olympus, Tokyo).

### SUPPLEMENTAL FIGURE LEGENDS

**Figure S1. Phylogenetic analysis of *Helicobacter* strains.**

A phylogenetic tree was constructed using the 23S rRNA gene sequences of 57 *Helicobacter* spp. strains, including 29 strains isolated from this study (13 strains of *H. suis* and 16 strains of *H. pylori*) and 28 reference strains of gastric *Helicobacter* species. Light blue and pink indicate *H. suis* and *H. pylori* strains, respectively. Numbers indicate a bootstrap percentage, and the scale bar indicates the number of base substitutions per site. Filled and open circles indicate strains isolated from this study and reference strains (NCBI reference sequences), respectively.

**Figure S2. ELISA for identification of *H. suis* and *H. pylori* infection using 1800-fold diluted sera.**

Serum specimens (n=101) were divided into four groups: an *H. suis*-infection group (n=20), an *H. pylori*-infection group (n=17), a group with eradication of *H. pylori* infection (n=20), and a group with neither *H. suis* nor *H. pylori* infection (n=44). (A) ELISA for detecting *H. suis* infection. The absorbances at 450 nm of ELISA in the *H. suis* infection, *H. pylori* infection, post-eradication of *H. pylori* infection and non-infection groups were 2.678±0.381, 0.707±0.775, 0.678±0.739, and 0.483±0.555, respectively. ****$P$<0.0001, the *H. suis* infection group vs. the *H. pylori* infection, the post-eradication of *H. pylori* infection, or the non-infection group. The red dotted line indicates the cut-off value. The bar represents the mean with SD. (B) ROC curve constructed from the *H. suis*-infection vs. the *H. suis* non-infection groups, i.e., the *H. pylori* infection, the post-eradication of *H. pylori* infection, and the non-infection group. Numbers in parentheses indicate the 95% CI. The red slanted line is the reference line. (C) ELISA for detecting *H. pylori* infection. The absorbance at 450 nm of ELISA in the *H. suis* infection group, *H. pylori* infection, post-eradication of *H. pylori* infection and non-infection groups were 0.903±0.640, 1.912±0.525, 1.243±0.837, and 0.580±0.618, respectively. ****$P$<0.0001, the *H. pylori* infection group vs. the *H. suis* infection or the non-infection group. #$P$>0.05, the *H. pylori* infection vs. the post-eradication of *H. pylori* infection group. The blue dotted line indicates the cut-off value. (D) ROC curve constructed from the *H. pylori* infection group vs. the *H. pylori* non-infection groups, including the *H. suis* infection and the non-infection groups. Numbers in parentheses indicate the 95% CI. The red slanted line is the reference line.

**Figure S3. Identification of *H. suis* and *H. pylori* in gastric biopsy specimens.**

Gastric mucosa sections of the antrum of specimens 04029 (A-C; *H. suis* infection) and 04220 (D-F; *H. pylori* infection) were stained with Giemsa (A and D), IHC using an anti-*H. pylori* monoclonal antibody (B and E), or IHC using an anti-*H. suis* polyclonal antibody (C and F). Arrows indicate microorganisms (A and D). Magnification, x450 and x1200 (inset). Scale bars, 25 µm and 10 µm (inset).
